# Effect of Temperature on the Transmission of COVID-19: A Machine Learning Case Study in Spain

**DOI:** 10.1101/2020.05.01.20087759

**Authors:** Amir Abdollahi, Maryam Rahbaralam

## Abstract

The novel coronavirus (COVID-19) has already spread to almost every country in the world and has infected over 3 million people. To understand the transmission mechanism of this highly contagious virus, it is necessary to study the potential factors, including meteorological conditions. Here, we present a machine learning approach to study the effect of temperature, humidity and wind speed on the number of infected people in the three most populous autonomous communities in Spain. We find that there is a moderate inverse correlation between temperature and the daily number of infections. This correlation manifests for temperatures recorded up to 6 days before the onset, which corresponds well to the known mean incubation period of COVID-19. We also show that the correlation for humidity and wind speed is not significant.

## 1. Introduction

Coronavirus disease 2019 (COVID-19) is an infectious disease first reported in Wuhan, China, in late December 2019. This disease is caused by severe acute respiratory syndrome coronavirus 2 (SARS-CoV-2) [1, 2], which has spread rapidly around the world. In response to this outbreak, the World Health Organization (WHO) characterised COVID-19 as a pandemic on 11 March [3]. As of today, the current COVID-19 outbreak situation has confirmed over 3 million cases and 208 thousand deaths globally [4]. Spain is one of the most affected countries with over 210 thousand confirmed cases and 23 thousand confirmed deaths [5] (data reported on April 28th 2020). COVID-19 infection could cause severe respiratory disorders with symptoms such as fever, coughing and shortness of breath. The average incubation period is 5-6 days with the longest incubation period of 14 days [6, 7, 8, 9, 10]. In addition to person-to-person transmission of COVID-19 through direct contact or droplets [11], studies have shown that geographical, and climatic factors are important factors in the transmission and survival of coronaviruses [12, 2, 13, 14, 15, 16, 17]. For example, temperature could increase or decrease the transmission risk by affecting the survival time of coronaviruses on surfaces [18, 19, 20, 21, 22]. Since the outbreak of COVID-19, many researchers have focused their efforts in understanding the influencing factors and transmission mechanism of this virus. These efforts are motivated by previous investigations on other viral infectious diseases such as SARS and Influenza. SARS is another coronavirus type that appeared in 2002 and caused a pandemic with more than 8 thousand infected people in 26 countries. Noticeable correlations between the SARS cases and the environmental temperature were reported [19, 23]. SARS transmission appeared to be dependent on seasonal temperature changes and the multiplicative effect of hospital infection [24]. It has also been documented that several factors, including climatic conditions (temperature and humidity), and population density might have affected the patterns of influenza spread and diversification [25].

Several studies have demonstrated that there is a correlation between meteorological factors, such as temperature and humidity, and the transmission of COVID-19 [26, 23]. Recent work has shown that there is a significant positive effect of the diurnal temperature range (DTR) on the daily mortality of COVID-19, and a significant negative association between COVID-19 mortality and ambient temperature as well as absolute humidity [27]. Tosepu and coworkers showed that only average temperature was significantly correlated with COVID-19 [28]. However, other studies have shown that temperature and humidity are not correlated with the transmission of COVID-19 [29, 30, 31]. This contradiction suggests a need for further investigation on this topic.

In this study, we explore the correlation between meteorological conditions and the daily increment of infected people due to COVID-19 in Spain. Given that Spain is the most climatically diverse country in Europe [32] and has one of the highest numbers of infected people, it becomes a potential case study country for this purpose. First, we introduce the methods used in this paper, including the study area, data collection, study period, and data analysis methods, then we present our analysis and results. Finally, we conclude the primary outcomes of our study.

## 2. Methods

### 2.1. Case study area

We consider the three most populous autonomous communities Anadalucía, Cataluña, and Madrid in Spain for our case study. Table 1 shows the population of these three communities in 2019 [33] and the number of confirmed COVID-19 cases until April 28th, 2020 [34]. In addition to their high population, these communities are located in different parts of Spain with varying conditions of climate. Figure 1 shows the average temperature of the three communities during March 2020. A temperature variation of up to about 16 °C is observed between these communities, highlighting the climate diversity of Spain.

**Table 1:** Population in 2019 [33] and the number of confirmed COVID-19 cases [34] in the three most populous autonomous communities in Spain until April 28th, 2020.

| Community | Population | Confirmed cases |
| --- | --- | --- |
| Anadalucía | 8,446,561 | 13,250 |
| Cataluña | 7,609,499 | 49,185 |
| Madrid | 6,685,471 | 63,989 |

**Figure 1:**
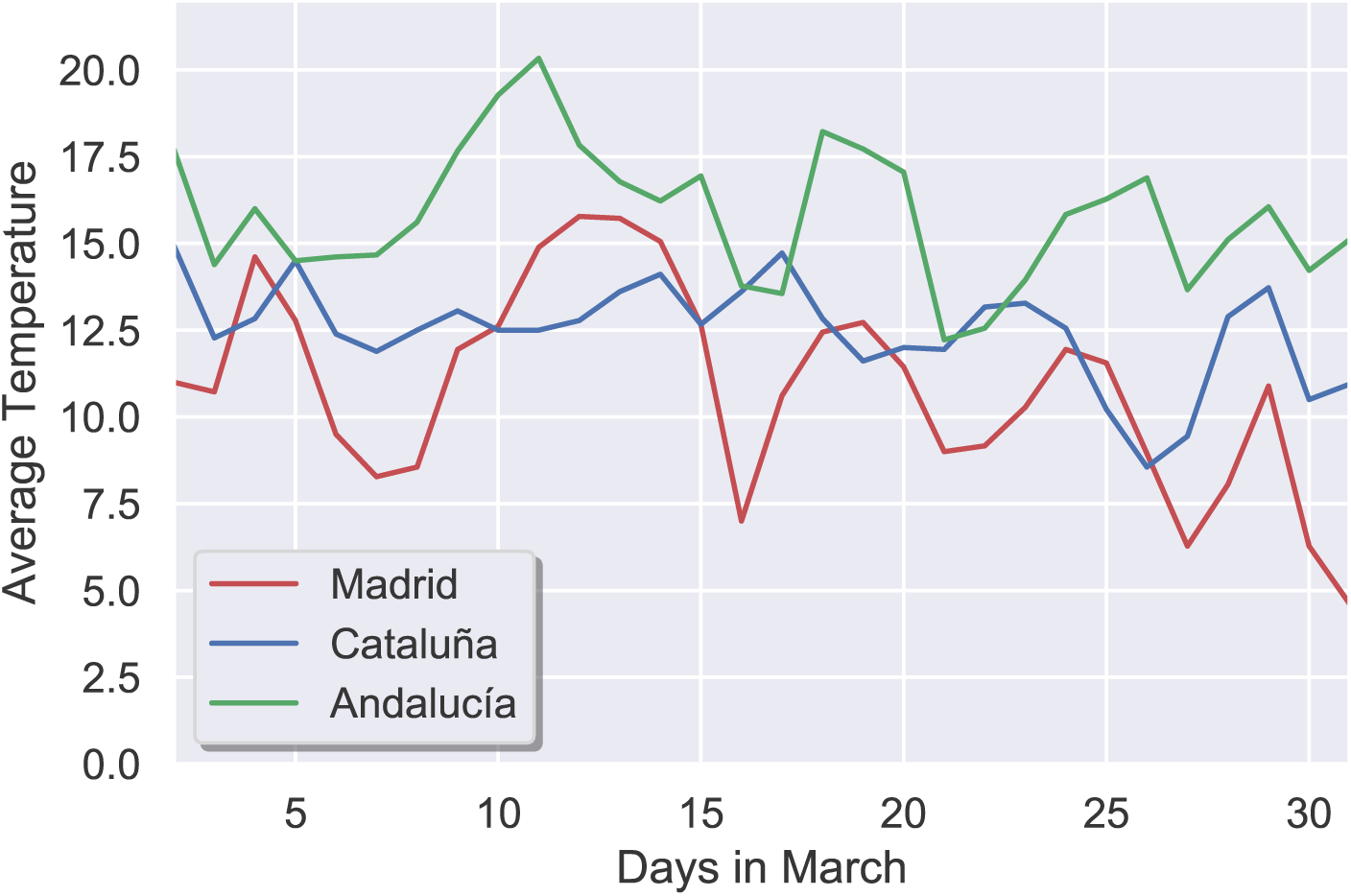
Average temperature during March 2020 in the three most populous autonomous communities in Spain; Madrid, Cataluña, and Andalucía.

### 2.2. Data collection

To study the effect of meteorological conditions on the transmission of COVID-19, here we consider the daily increment of infected people in the three communities. This dataset was extracted from Datadista Github repository [34], containing the updated daily information according to the calendar and publication of the Spanish Ministry of Health. For each daily record, we also obtain meteorological data such as the averages of temperature, humidity, and wind speed for the same day and the previous 13 days, leading to a total of 42 predictors. These data were extracted from Wunderground, an online open-source weather database [35]. After merging these predictors with the daily increment of infected people for each community, then we concatenate the three databases to form a single dataset for our analysis.

### 2.3. Case study period

Figure 2 shows the daily increment of infected people for the communities of Madrid, Cataluña, and Anadalucía from March 1st until April 28th, 2020. All the three communities reached the peak of daily infections until the end of March with an upward trend during the month. A state of alarm and a national lockdown was imposed on March 14th [36], whose effect manifests as a downward trend in the number of daily cases from the beginning of April (day 31). To minimise the impacts of the lockdown on our analysis, we consider the upward transmission period during March, see Fig 2. Also, we focus our case study on the last 17 days of March to capture all the possible transmissions during the first 14 days, known as the maximum incubation period for the virus [8, 9, 10].

**Figure 2:**
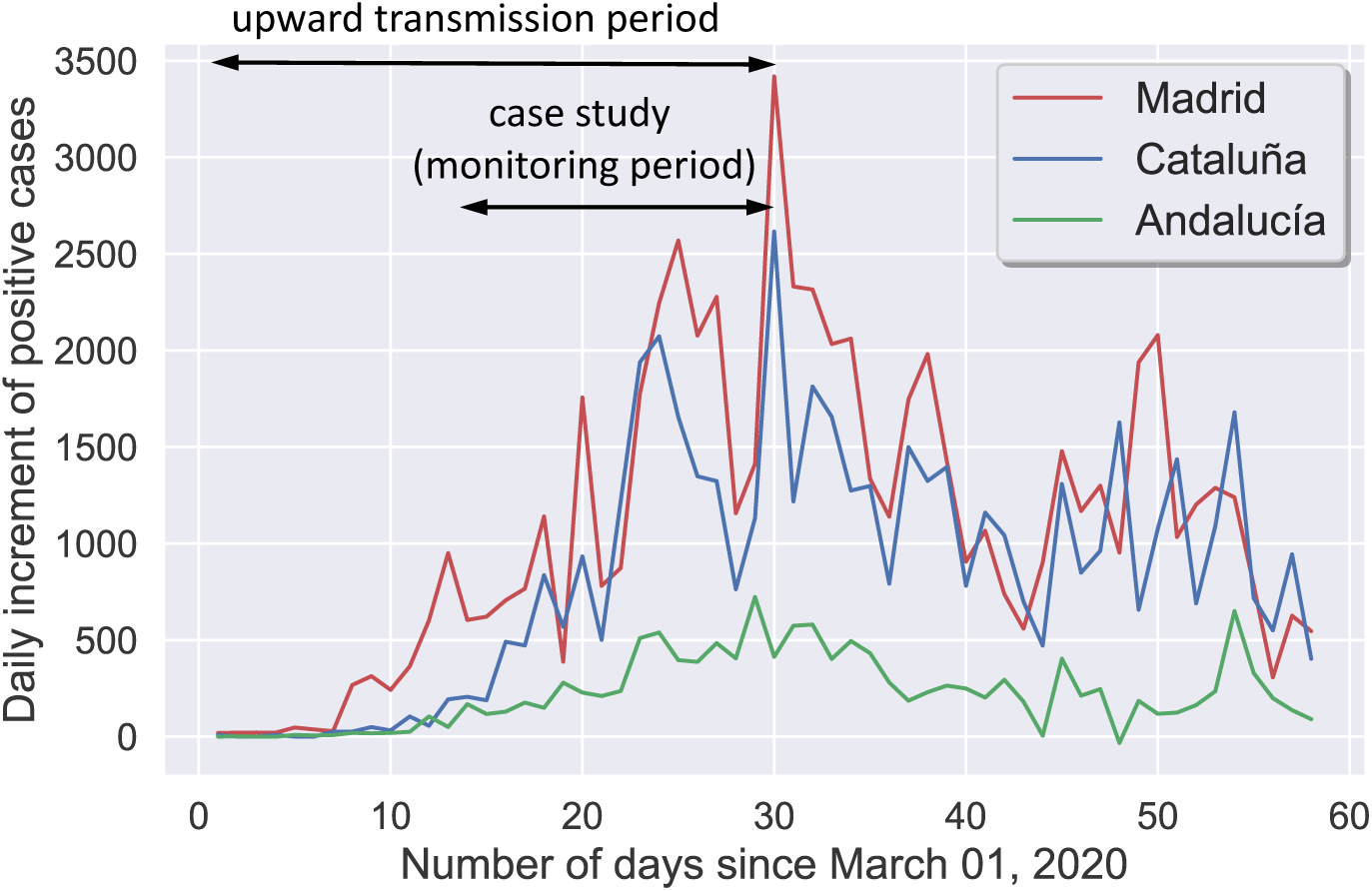
Daily increment of infected people as a function of the number of days since March 1st, 2020, for the three most populous autonomous communities in Spain; Madrid, Cataluña, and Andalucía. For our case study, we consider a monitoring period of 17 days when the number of infections shows an upward trend.

### 2.4. Data analysis

In this study, we use the Pearson’s correlation coefficient as a measure of the linear association between the predictors and the target. Like other correlation coefficients, the Pearson’s correlation varies between −1 and +1 with 0 implying no correlation. The p-value roughly indicates the probability of an uncorrelated system producing datasets that have a Pearson correlation at least as extreme as the one computed from these datasets. We also employ linear regression, a supervised machine learning technique, to find the relationship between the most important predictors and the target [37]. Here, we use Ordinary Least Squares (OLS) to estimate the unknown regression coefficients. A range of plausible values for these coefficients can be obtained by calculating confidence intervals.

## 3. Results and Discussion

Figure 3 presents the correlation between the meteorological predictors and the daily increment of infected people (target). It is interesting to observe that the averages of temperature up to the previous 6 days (T-AVG, and T-AVG-1 to −6) show a moderate negative correlation range between −0.7 to −0.5 (p-value < 10^−3^). This result corresponds well with the mean incubation period of COVID-19, which was estimated to be 5-6 days [8, 9, 10]. We note that none of the 14 temperature predictors shows a positive correlation with the target. Humidity and wind speed do not indicate this consistency, and they also show a weak correlation in the range of −0.5 to 0.5.

**Figure 3:**
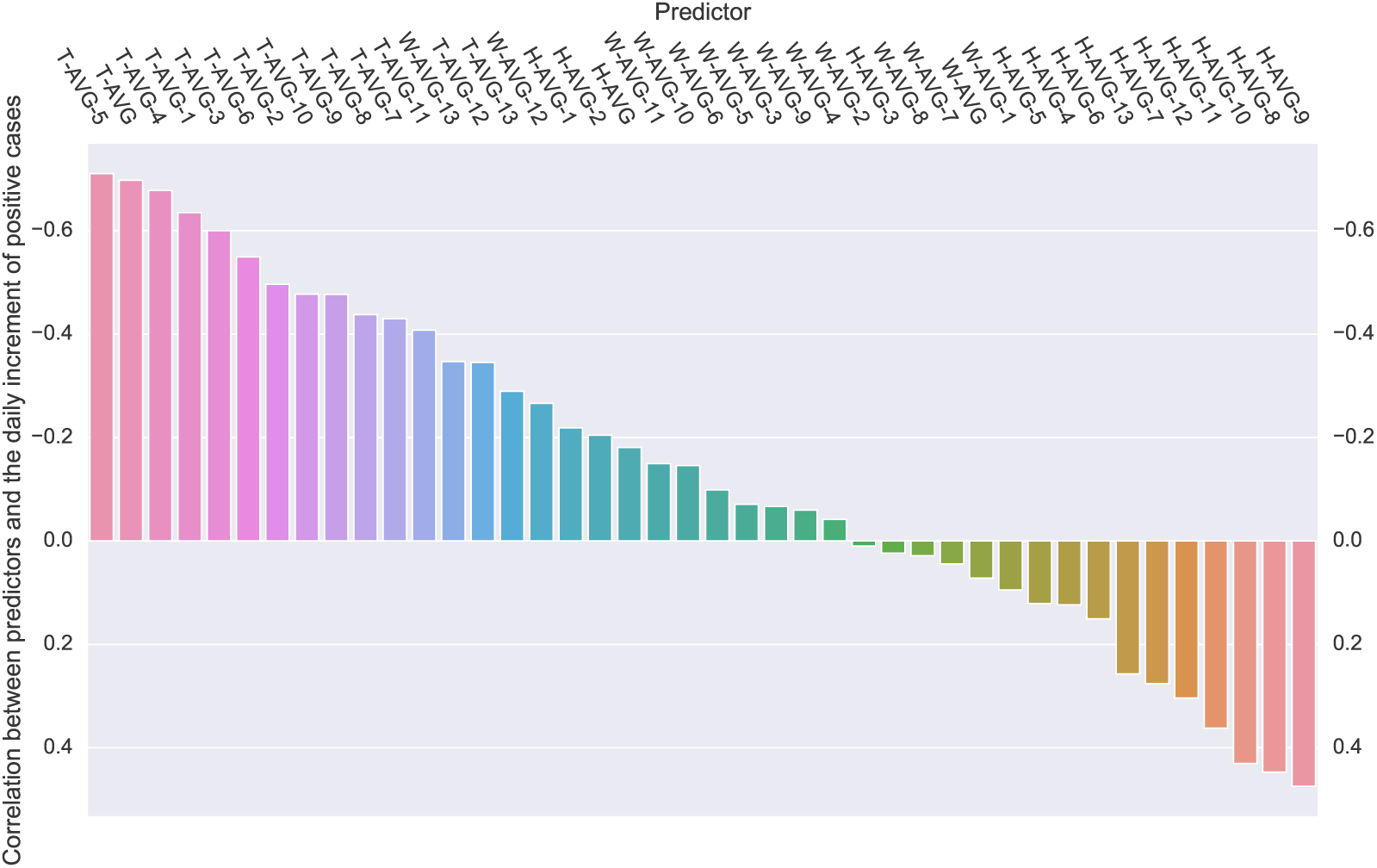
Correlation between the meteorological predictors and the daily increment of infected people. The averages of temperature, humidity and wind speed are represented by T-AVG, H-AVG, and W-AVG, respectively. The following number shows the number of days’ lag between the date when these meteorological predictors were measured and the date when the number of infections is considered.

We perform linear regression between the target and the 5-day lag temperature (T-AVG-5), which shows the highest correlation. Figure 4 shows the data points, the regression line, as well as the confidence interval (95%). Considering a linear regression model *I* = *β*_0_ + *β_1_T*, *I* and *T* being the daily increment of infected people and the average temperature, respectively, an intercept of *β*_0_ = 3556 (95% CI, 2813 to 4298) and a slope of *β*_1_ = −194 ^°^C^-1^ (95% CI, −248 to −140) are obtained from the linear regression results. In other words, a 1°C increase in average temperature leads to a decrease of 194 daily positive cases (95% CI, 140 to 248). We also perform linear regression considering the other important temperature predictors. Table 2 summarizes the regression coefficients. The coefficient of *β*_1_ shows a range from −178 ^°^C^-1^ to −136 ^°^C^-1^, while the intercept *β*_0_ varies from 2691 to 3556 among different predictors.

**Figure 4:**
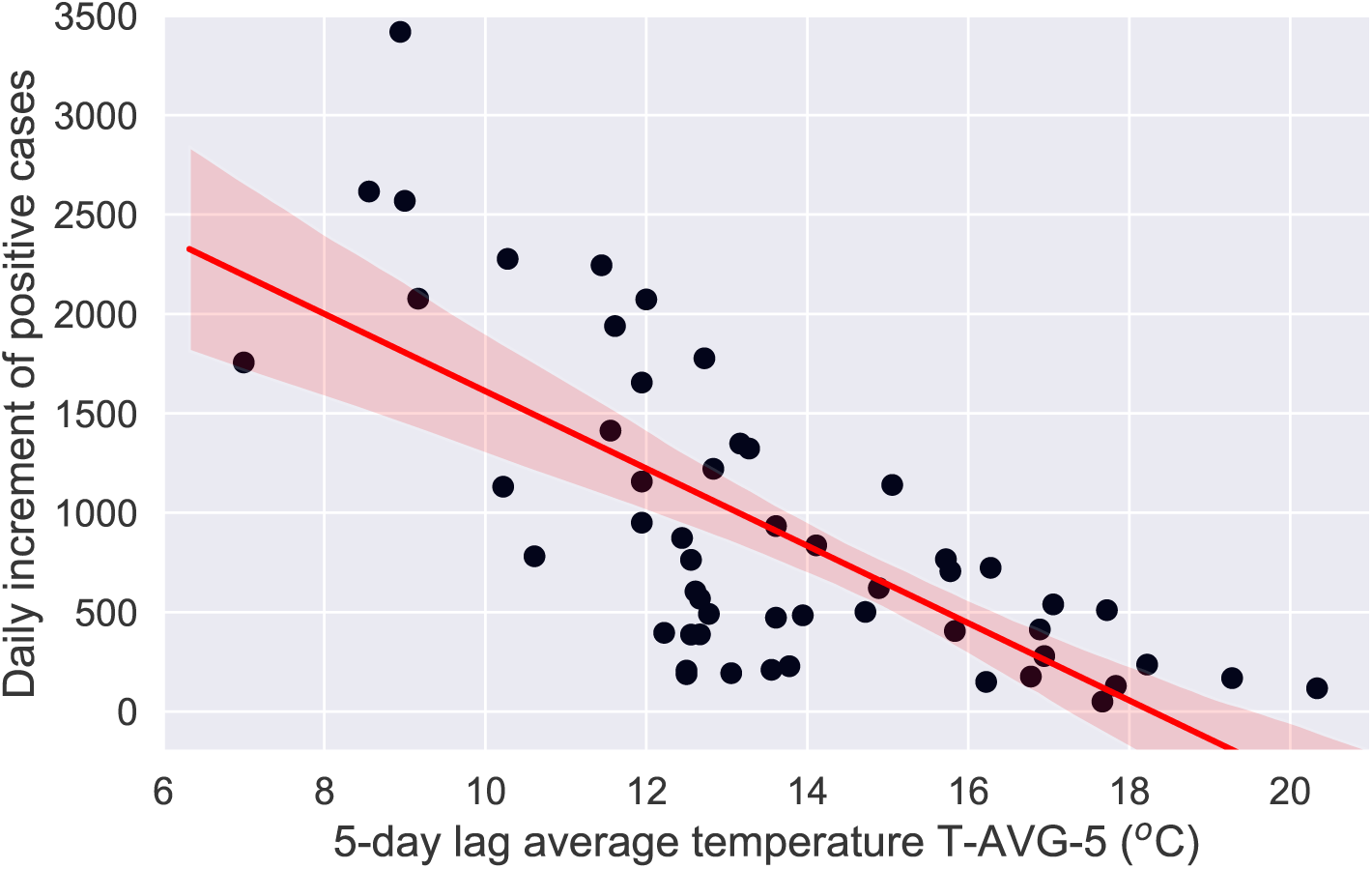
Daily increment of infected people as a function of the 5-day lag average temperature (T-AVG-5). The regression line and the confidence interval (95%) are highlighted in red.

**Table 2:**
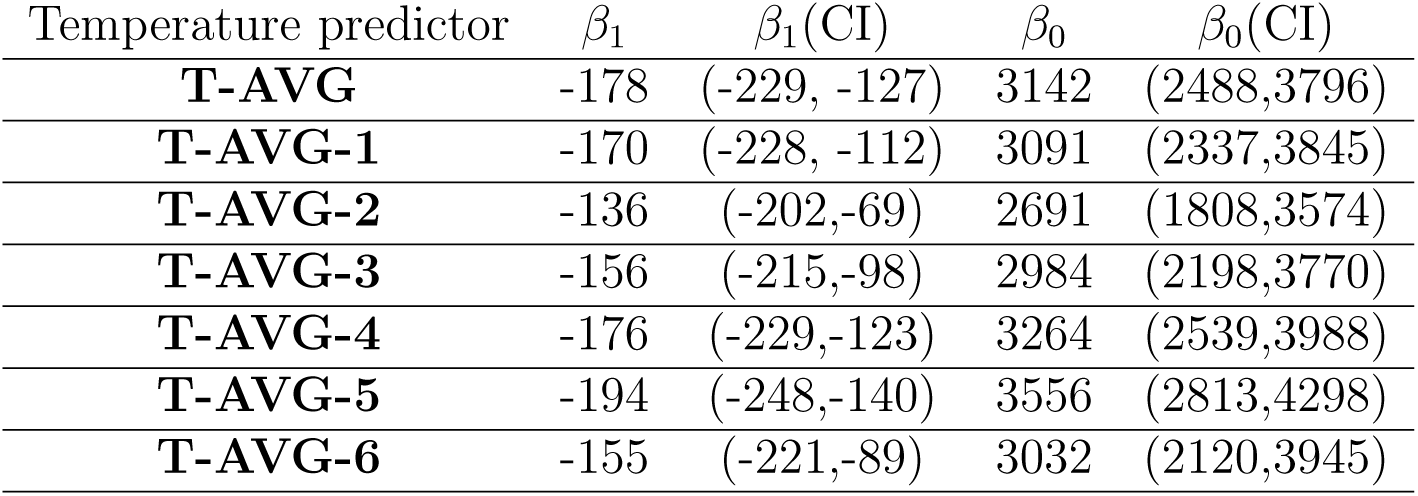
Regression coefficients and 95% confidence interval (CI) for important temperature predictors.

| Temperature predictor | $\beta_1$ | $\beta_1(\text{CI})$ | $\beta_0$ | $\beta_0(\text{CI})$ |
| --- | --- | --- | --- | --- |
| **T-AVG** | -178 | (-229, -127) | 3142 | (2488,3796) |
| **T-AVG-1** | -170 | (-228, -112) | 3091 | (2337,3845) |
| **T-AVG-2** | -136 | (-202,-69) | 2691 | (1808,3574) |
| **T-AVG-3** | -156 | (-215,-98) | 2984 | (2198,3770) |
| **T-AVG-4** | -176 | (-229,-123) | 3264 | (2539,3988) |
| **T-AVG-5** | -194 | (-248,-140) | 3556 | (2813,4298) |
| **T-AVG-6** | -155 | (-221,-89) | 3032 | (2120,3945) |

## 4. Conclusions

We have presented a machine learning case study to evaluate the effect of meteorological factors, such as temperature, humidity, and wind speed on the transmission of the novel coronavirus (COVID-19). We have found that the average temperatures of up to 6 previous days have a moderate negative correlation with the daily increment of infected people. This result is relevant since the mean incubation period of COVID-19 is known to be around 5-6 days [8]. We have also performed a linear regression which has shown a decrease of about 200 daily infected people per 1°C increase in average temperature. We note that due to several uncertainties involved in testing and collecting the number of infected people, and other factors affecting the transmission, our analysis and results need to be interpreted with caution. However, our results could provide a clue to understanding the transmission mechanism of COVID-19.

## Data Availability

The datasets used and/or analyzed during the current study are available from the websites.

https://github.com/datadista/datasets/tree/master/COVID%2019

https://www.wunderground.com

